# The resurgence risk of COVID-19 in the presence of immunity waning and ADE effect: a mathematical modelling study

**DOI:** 10.1101/2021.08.25.21262601

**Authors:** Weike Zhou, Biao Tang, Yao Bai, Yiming Shao, Yanni Xiao, Sanyi Tang

**Author notes:** Corresponding author *Email addresses:* (Yanni Xiao), (Sanyi Tang). Equal Contributor (Weike Zhou, Biao Tang, Yao Bai).

## Abstract

Since the end of 2020, the mass vaccination has been actively promoted and seemed to be effective to bring the COVID-19 pandemic under control. However, the fact of immunity waning and the possible existence of antibody-dependent enhancement (ADE) make the situation uncertain. We developed a dynamic model of COVID-19 incorporating vaccination and immunity waning, which was calibrated by using the data of accumulative vaccine doses administered and the COVID-19 epidemic in 2020 in mainland China. We explored how long the current vaccination program can prevent China in a low risk of resurgence, and how ADE affects the long-term trajectory of COVID-19 epidemics. The prediction suggests that the vaccination coverage with at least one dose reach 95.87%, and with two-doses reach 77.92% on August 31, 2021. However, even with the mass vaccination, randomly introducing infected cases in the post-vaccination period can result in large outbreaks quickly in the presence of immunity waning, particularly for SARS-CoV-2 variants with higher transmission ability. The results showed that with the current vaccination program and a proportion of 50% population wearing masks, mainland China can be protected in a low risk of resurgence till 2023/01/18. However, ADE effect and higher transmission ability for variants would significantly shorten the protective period for more than 1 year. Furthermore, intermittent outbreaks can occur while the peak values of the subsequential outbreaks are decreasing, meaning that subsequential outbreaks boosted the immunity in the population level, which further indicating that catching-up vaccination program can help to mitigate the possible outbreaks, even avoid the outbreaks. The findings reveal that integrated effects of multiple factors, including immunity waning, ADE, relaxed interventions, and higher transmission ability of variants, make the control of COVID-19 much more difficult. We should get ready for a long struggle with COVID-19, and should not totally rely on COVID-19 vaccine.

## 1. Introduction

Vaccination against COVID-19 has been regarded as an important measure to break the transmission chain of SARS-CoV-2 infections. Several vaccines against SARS-CoV-2 have been developed and approved by the WHO since the end of the year 2020 [1]. In mainland China, the two-doses vaccination program has been actively and widely promoted by injecting the inactivated vaccines. The vaccination of high risk populations was initiated on December 15, 2020, and over 1.82 billion COVID-19 vaccination doses had been administered by August 13, 2021 [2]. Citing the real epidemic data [3], the mass vaccination strategy seems to be able to stop the COVID-19 pandemic. However, many emerging evidences indicate that the vaccination will not help to eradicate the spread of SARS-CoV-2. On one hand, the immunity waning and the limited efficacy of the vaccines result in a large number of the vaccinated population still susceptible to SARS-CoV-2, particularly in terms of SARS-CoV-2 variants [4–6]. On the other hand, the existence of antibody-dependent enhancement (ADE) in the infection of SARS-CoV-2 has been reported recently [7].

ADE is usually referred to the phenomenon in which the pre-existing antibodies enhance the infectivity of a secondary infected virus, and facilitate the transmission of the virus. ADE phenomenon is well documented between different dengue serotypes [8–10] and Zika virus [11–13], and the infection of other corona viruses, including MERS [14] and SARS [15]. In a recent publication, Liu et al. showed that COVID-19 patients can not only produce antibodies against the RBD of the spike protein to block the SARS-CoV-2 infections, but also produce the anti-spike antibodies enhancing ACE2 binding, consequently enhancing the infectivity of SARS-CoV-2 [7]. This supports the existence of the ADE in SARS-CoV-2 infections. In [16], the author concluded two possible ways to induce the ADE effect by COVID-19 vaccines. Lots of mathematical models have been developed to discuss the impact of ADE between different dengue serotypes [17–19], or between dengue and Zika [20–22], on their transmission dynamics and viral dynamics. During the very early stage of the development of COVID-19 vaccines, several researchers posted their concerns on that ADE can be a potential safety issue [23, 24]. However, it remains unclear and challenging how the ADE effect in SARS-CoV-2 infection will affect the trajectory of the COVID-19 pandemic when we use COVID-19 vaccines in the population.

What’s more, Choe et al. conducted a clinical study to measure the changes of neutralizing antibodies in both symptomatic and asymptomatic SARS-CoV-2 infection, and found that the geometric mean titer of neutralizing antibodies declined from 219.4 at 2 months to 143.7 at 5 months after infection [5]. Similarly, in [6], based on a longitudinal study of 517 COVID-19 patients, the authors observed different levels of immunity waning after symptoms onset. The immunity waning makes the prospect of achieving herd immunity increasingly remote, that is, the prominence of herd immunity being touted as a solution to the pandemic might be about to change [25]. Therefore, it’s urgent to evaluate the impact of the immunity waning on the trends of COVID-19 epidemics, and it’s essential to re-design the optimal control interventions in combating with COVID-19 pandemic in a long-term. This also remains challenging.

Hence, the immunity waning and ADE make the long-term trajectory of COVID-19 epidemics be full with uncertainty. This study aims to develop a mathematical model describing the transmission process of COVID-19 and the two-doses vaccination program incorporating with the immunity waning and ADE effect, to investigate the effect of immunity waning and ADE. We used the COVID-19 epidemic data from January 23 to April 8, 2020 and cumulative vaccine doses administered in mainland China to inform model parameters and conducted sensitivity analysis to evaluate how long the program can protect China in a low risk of resurgence, and how ADE will affect the transmission dynamics of COVID-19 epidemics. Findings from this study will provide important information for policymakers on the critical time of implementing strict control measures and when the catch-up vaccination program should be launched.

## 2. Methods

We proposed a modelling framework which incorporates the infection and transmission process of COVID-19 with the two-doses vaccination program and the immunity waning. The model can be reduced to a transmission model only and a vaccination model only to estimate the parameters related with the transmission and the vaccination by using epidemic data and vaccination data. The full model was used to simulate different scenarios during the post-pandemic period to evaluate the resurgence risk and explore the low risk protective period.

### 2.1. Model overview

We developed a dynamic model of COVID-19 infection and transmission incorporating with the vaccination program and immunity waning in mainland China (Fig. 1 and model (1) in SI). The modelling framework *SEIARS* is used to describe the transmission dynamics, where the route that recovered individuals *R* can go back to susceptible population *S* is used for modelling the immunity waning. The population is further divided into three categories according to their vaccination states: not vaccinated, vaccinated by one-dose, vaccinated by two-doses. Only part of the vaccinated population is effectively protected, hence the vaccinated but not effectively protected population also evolve a transmission dynamic with the same structure of *SEIARS*. Similar to the recovered population, individuals either effectively protected by the first dose or by two-doses vaccine will also lose their immunity after a period of time. It should be mentioned that the recovered population have the same level of immunity as those effectively protected by two-doses. Given the possibility of the existence of ADE, we assumed that the susceptibility of the individuals lost their immunity is higher than those without any immunity before. In particular, *κ* is denoted as the modification factor of the susceptibility. The detailed assumptions and the corresponding model equations are shown in SI.

**Figure 1:**
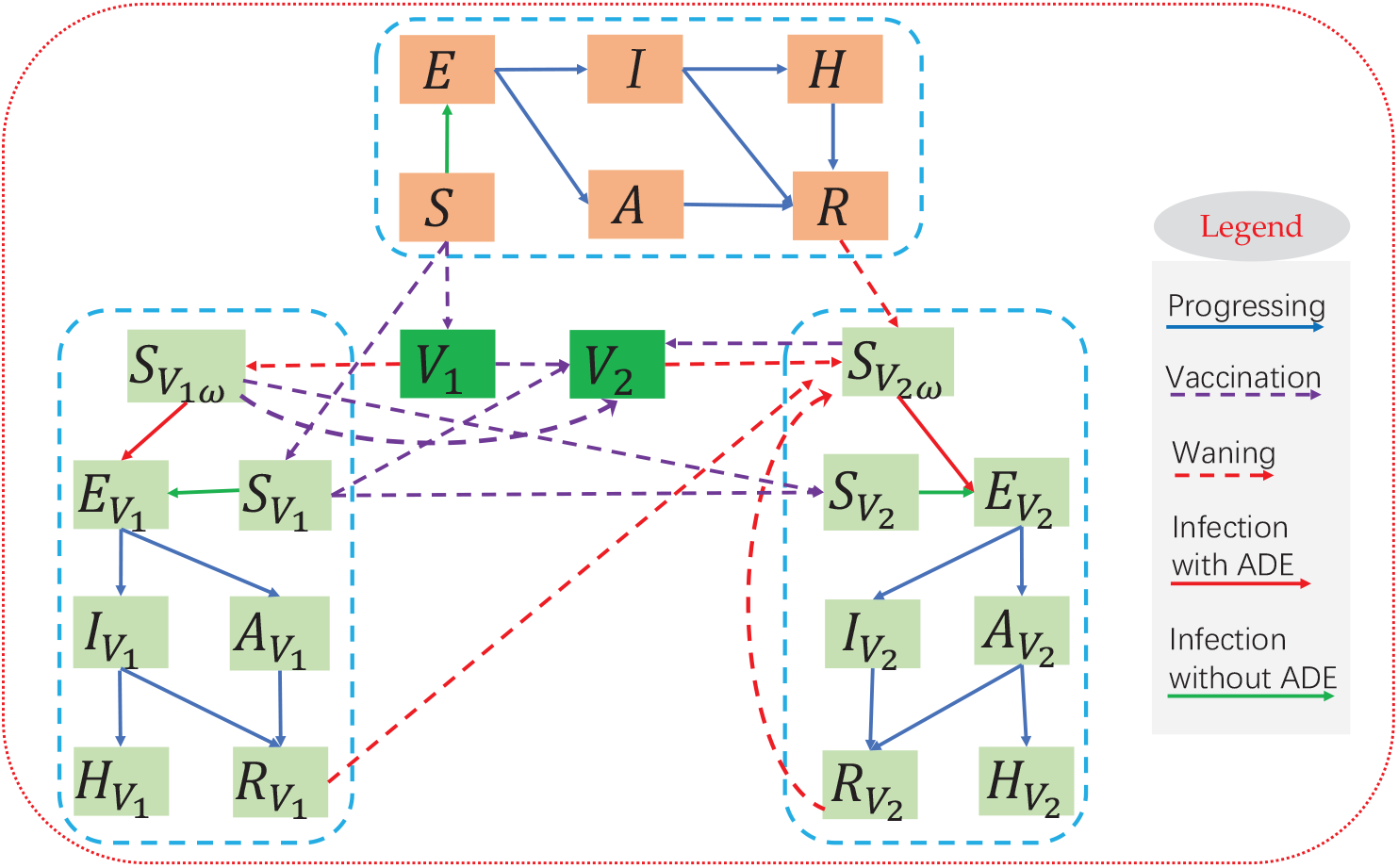
Schematic diagram to illustrate the transmission of COVID-19 incorporating with the vaccination program and immunity waning.

### 2.2. Data

We obtained the data of the COVID-19 epidemic, and the data of the mass vaccination program in mainland China from the National Health Commission of the People’s Republic of China [2] and Our World in Data [26]. The epidemic data includes the number of daily confirmed cases and deaths from January 23, 2020 to April 8, 2020 in mainland China, as shown in Fig. 2 (a) and 2 (b). The data of the vaccination program, collected from December 15, 2020 to June 29, 2021, contains the cumulative vaccine doses administered and the daily vaccine doses administered in mainland China, see Fig. 2 (c) and 2 (d). It should be mentioned that the vaccination data is available from December 15, 2020 with a report of accumulative 1,500,000 doses used in the population. However, during the period from December 15, 2020 to March 23, 2021, the data was not released daily, hence is intermittent. Since March 23, 2021, the vaccination data was reported per day.

**Figure 2:**
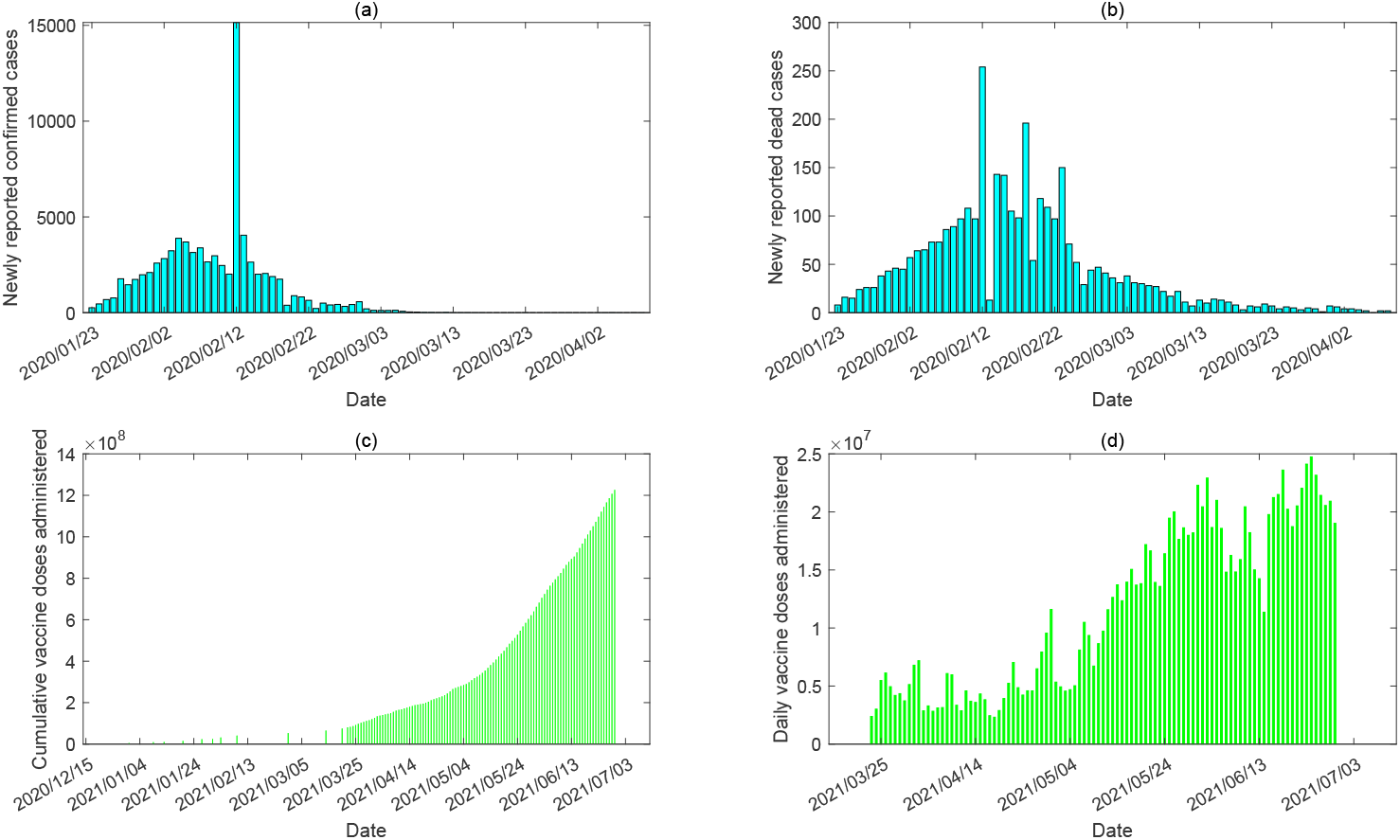
The epidemic data of COVID-19 in mainland China from January 23, 2020 to April 8, 2020 ((a)-(b)), and the vaccine doses administered in mainland China from December 15, 2020 to June 29, 2021 ((c)-(d)). (a) Daily reported confirmed cases; (b) Daily reported dead cases; (c) Cumulative vaccine doses administered; (d) Daily vaccine doses administered.

### 2.3. Model calibration

To calibrate the model, we firstly use the data of the COVID-19 epidemic from January 23, 2020 to April 8, 2020 in mainland China to estimate the parameters related to transmission dynamics. As there is no vaccination during this period, the full model can be reduced to a model without vaccination (i.e. model (3) in SI). Note that, because the epidemic lasted less than four months and only a very small proportion of the whole population was infected, we didn’t consider the immunity waning in the reduced model. Considering the continuously enhanced control interventions implemented by the government, we introduced a time-dependent transmission rate and diagnose rate. Based on these assumptions, we fit model (3) to the epidemic data of mainland China in 2020. To this end, we first fixed several parameters from the literature and initial values from the database, as listed in Table 1. Given the randomness of reported cases, we used a bootstrap method to generate 1000 time series of daily confirmed cases and deaths from a Poisson distribution with mean given by the reported data, hence we obtained 1000 data set. We then use the least square method to fit the model to each data set.

**Table 1:**
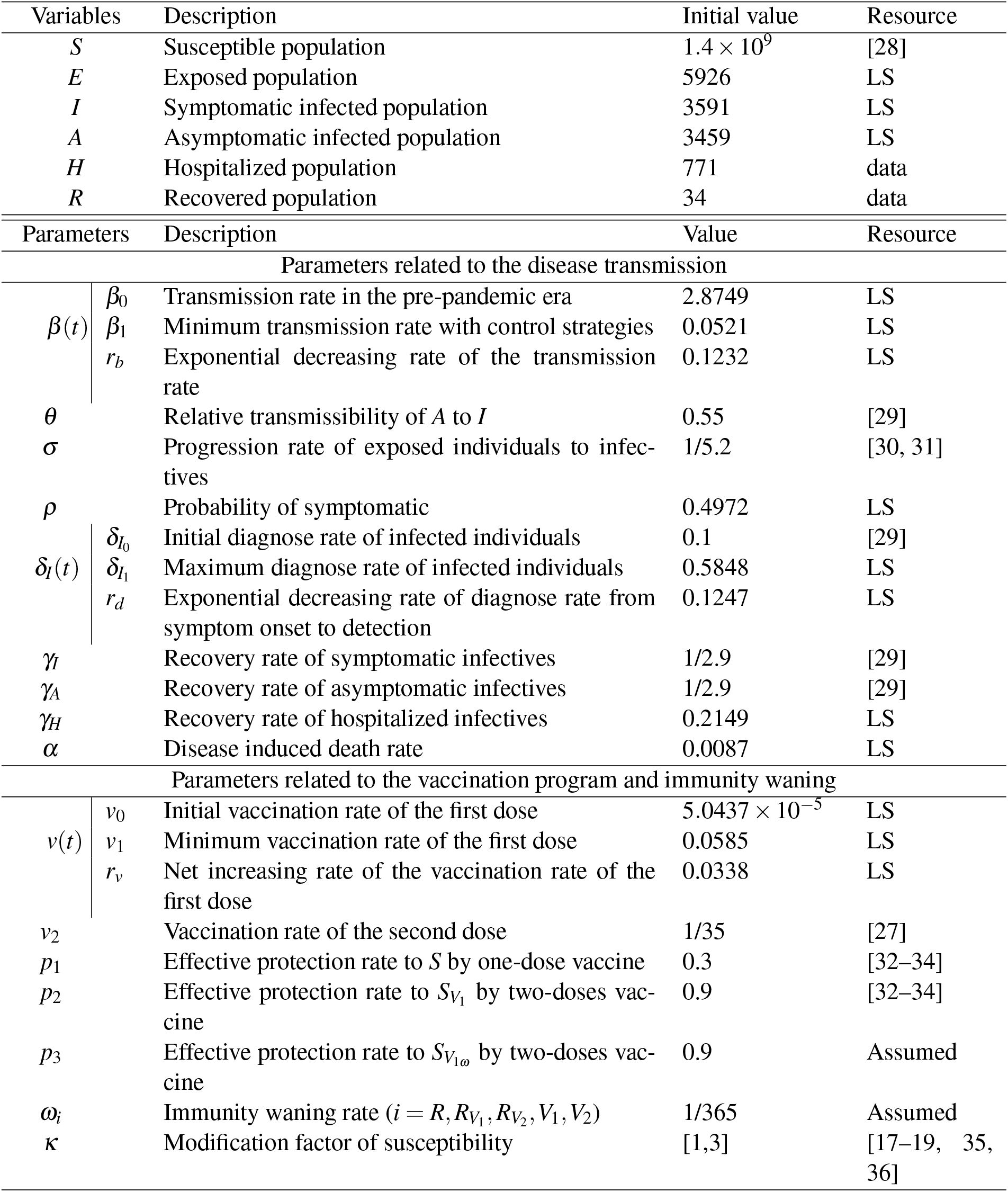
Definitions and values of variables and parameters

We further used the vaccination data in mainland China from December 15, 2020 to June 29, 2021 to estimate the parameters related to the mass vaccination program in China. To this end, we deduced a vaccination dynamical model (model (6) in SI) from the full modelling framework. Considering the initial accessability of vaccines and the capacity of daily vaccination population, we set the rate, at which the population receive the first dose, as a logistic function of time. In contrast, the rate at which the individuals get the second dose is fixed as a constant. Note that the second doses is requested to be vaccinated in 3-8 weeks in China [27], hence we set 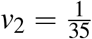. Based on the vaccination data, the initial time is set as December 15, 2020, and the initial conditions are *S*(0) = 1, 400, 000, 000, *S*_1_(0) = 1, 500, 000, *S*_2_(0) = 0. Then, by using the similar methods as in fitting the epidemic data, we fitted the vaccination dynamic model to 1000 vaccination data set.

### 2.4. Parameter setting for simulations

Because of the highly improved testing capacity, we set the diagnose rate as the estimated maximum rate (i.e. 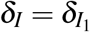) when carrying the simulations. The baseline protective rate of the first dose and the second dose vaccine are *p*_1_ = 0.3, *p*_2_ = 0.9, respectively [32–34]. Suppose the immunity produced by infection or vaccination lasts 1 year on average, then the immunity waning rate 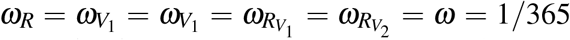 per day. Using face mask is recognized as an useful self-protective way to prevent the COVID-19 infection. During the COVID-19 epidemic in 2020 in China, the proportion of people wearing face masks increase from 0% to almost 100%, thus face mask use would be a normalized prevention and control intervention. Assume that the baseline proportion of face mask use is around 50% in the post-pandemic era, and the effectiveness of face mask in preventing COVID-19 infection or infecting others is 80% based on a recent meta-analysis on the effectiveness of face mask use [37]. Thus the baseline transmission rate with a normalized control intervention of wearing masks would be (1 − 50% × 80%)*β*_0_ = 60%*β*_0_. Given the enhanced intervention, the transmission rate can decrease further, and considering the higher transmission ability of SARS-CoV-2 variants, the transmission rate can also be higher than the initial value *β*_0_. Consequently, when performing sensitivity analysis, we choose the transmission rate from 0.4*β*_0_ to 1.5*β*_0_. In the absent of real data, we pick up a range of [1, 3] for the modification factor of ADE (*κ*) from the studies on the ADE in dengue infections [17–19, 35, 36].

## 3. Main results

### 3.1. Estimation results

The fitting results of the transmission dynamic model (model (3) with (4) and (5) in SI) to the epidemic data of mainland China in 2020 are shown in Fig. 3, with the best fitting curves marked as black. Based on the fitting results, we obtained the estimation of the unknown parameters (listed in Table 1), and also the estimated effective reproduction number (Fig. 3(c)). The fitting results of the vaccination dynamic model (model (6) with (7) in SI) to the vaccination data are shown in Fig. 4. And we obtained the estimation of the vaccination rates, as listed in Table 1. The estimation showed that the population vaccinated with at least one dose vaccine reached 56.4% (95% CI [55.38%, 57.08%]) while the population vaccinated with both two doses reached 32.02% (95%CI [31.93%, 32.06%]) on June 29, 2021 (the last data point). A further prediction shows that the population vaccinated with at least one dose can reach 95.87% (95%CI [91.12%,98.16%]) and people fully vaccinated reaches 77.92% (95% CI[73.33%,79.33%]) on August 31, 2021. As we can see, the vaccination coverage in China is very high till August 31, 2021, hence we assume to stop the routine vaccination program on August 31, 2021. Of course, we should complete the second dose injection for individuals who has been given the first dose already. Note that, unless otherwise stated, the considered time period is always to the end of year 2022 when performing the simulations.

**Figure 3:**
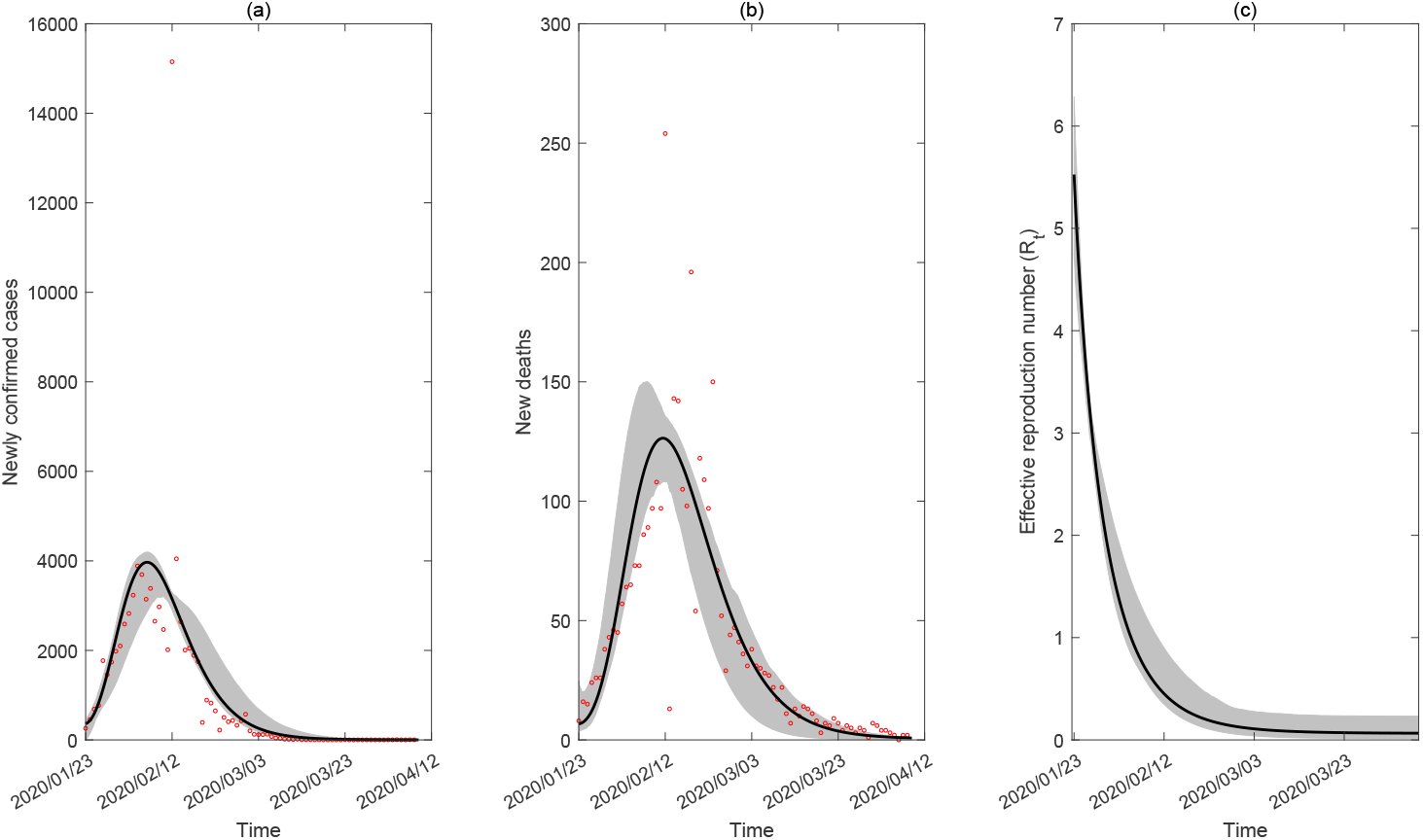
Model fitting results for the transmission dynamic model in mainland China in 2020. (a) The daily reported confirmed cases. (b) The daily reported dead cases. (c) The estimated effective reproduction number. The black curves are the estimated curves with the shadow areas as the corresponding 95% confidence intervals. The red circles in (a) and (b) are the observed data of the daily reported confirmed cases and the daily reported dead case from January 23, 2020 to April 8, 2020 in mainland China.

**Figure 4:**
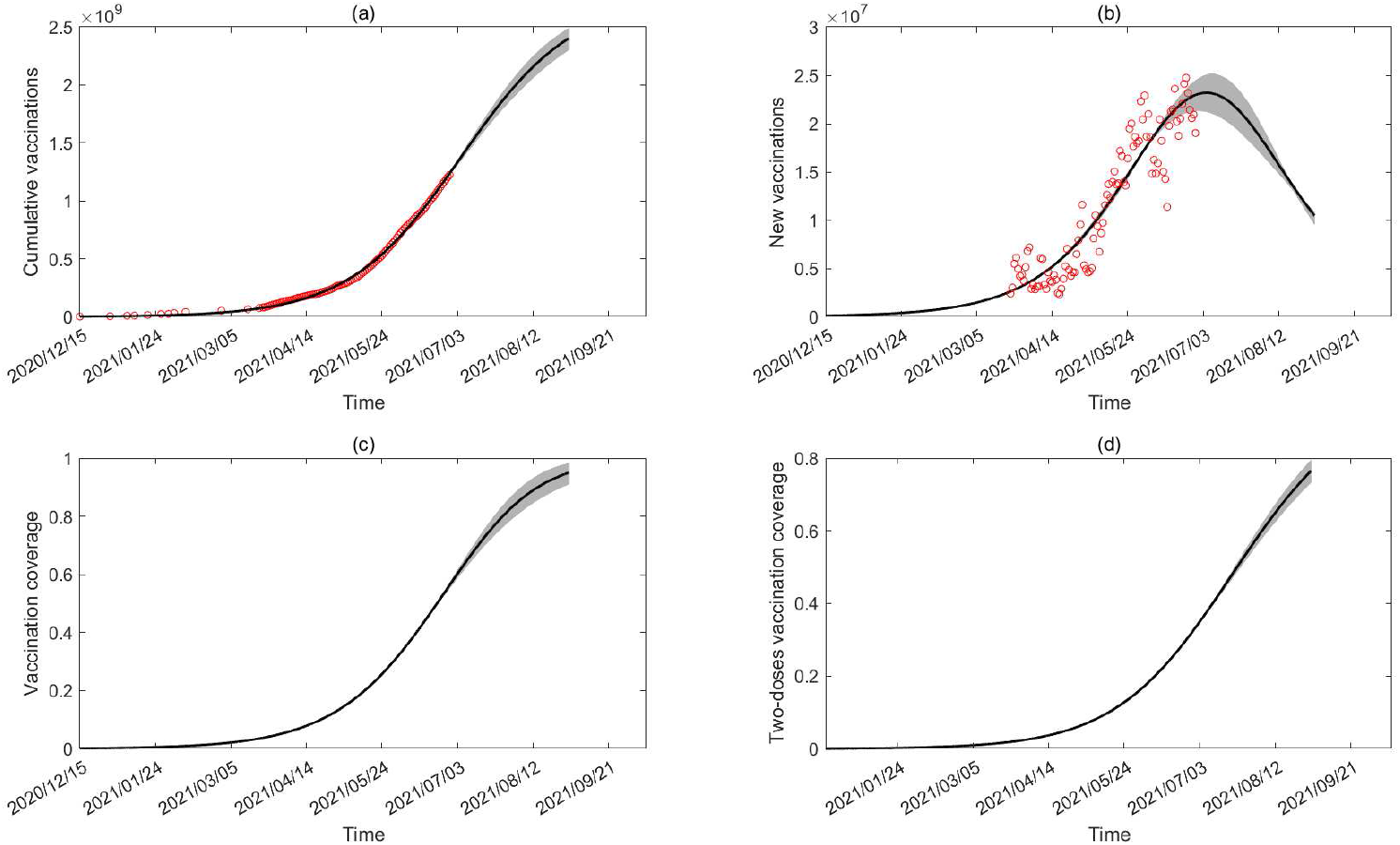
Model fitting results for the vaccination dynamic model. (a) The accumulative number of vaccine doses administered. (2) The daily number of vaccine doses administered. (c) Proportion of population vaccinated with at least one dose. (d) Proportion of population vaccinated with both two doses. The black curves are the estimated curves with the shadow areas as the corresponding 95% confidence intervals. The red circles in (a) and (b) are the reported data of the accumulative vaccine doses and daily vaccine doses administered from December 15, 2020 to June 29, 2021 in mainland China.

### 3.2. Resurgence risk evaluation

Based on the above estimation results, through numerical simulations, we focused on discussing the impact of immunity waning and ADE effect on the transmission dynamics of COVID-19, and evaluating the resurgence risk of COVID-19 in China. As we know, the COVID-19 epidemic in China is almost under control. The biggest challenge is that the imported cases have caused several local outbreaks in China. Therefore, we analyzed if there can be an large outbreak only with the mass vaccination or the vaccination plus a normalized control intervention by wearing masks, by randomly introducing several infected cases into the community.

Assume that 10 infected cases are introduced on September 1, 2021, Fig. 5 shows the number of newly confirmed cases and the effective reproduction number *R*_*t*_ during the transmission process, with different transmission rate and various ADE degree. It follows from Fig. 5(a) and 5(c) that, even without ADE (*κ* = 1), introducing infected cases would cause the large outbreaks (black curves) as the immunity waning. Worse still, ADE would facilitate the outbreak by bringing forward the peak time and increasing the peak value. Comparing Fig. 5(a) and Fig. 5(c), we find that the normalized intervention (*β* = 0.6*β*_0_) cann help to delay the outbreak and reduce the peak value. We also observed several subsequential epidemic waves in Fig. 5(a) and 5(c), while the peak values of the subsequential waves are decreasing. Furthermore, the ADE effect and the higher transmission ability can also increase the frequency of the outbreaks. Correspondingly, the effective reproduction number fluctuates around the threshold of unit, as shown in Fig. 5(b) and 5(d).

**Figure 5:**
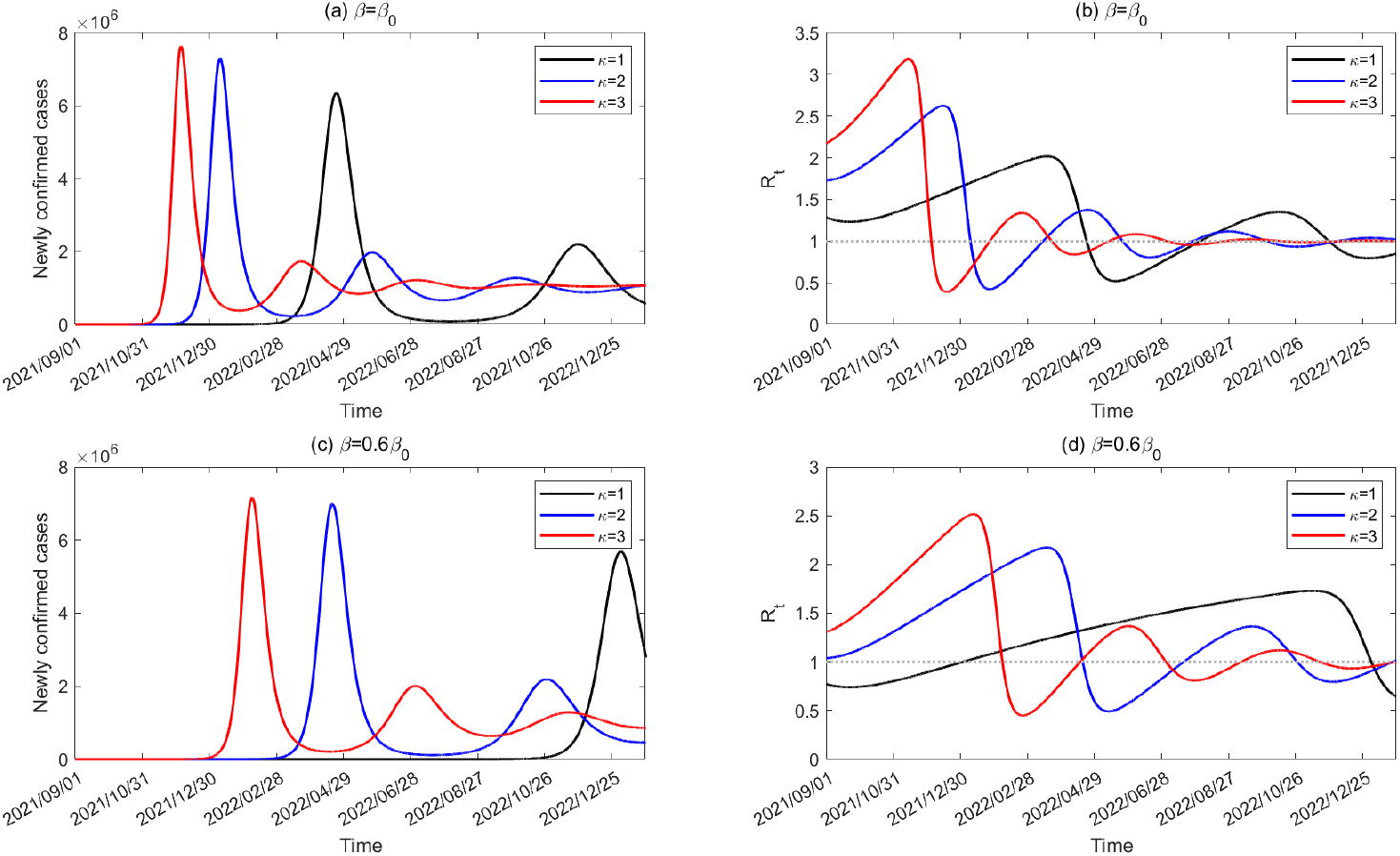
Effect of the ADE and the control interventions on the number of newly confirmed cases and effective reproduction number during the transmission process when introducing 10 infected cases on September 1, 2021.

In Fig. 5, we always set the introducing time as September 1, 2021, we then explored the impact of the introducing time on the transmission dynamics of COVID-19. To this end, by assuming the introducing time to be September 1, 2021, November 1, 2021, January 1, 2022, respectively, we further plotted the number of newly confirmed cases and effective reproduction number with a normalized control intervention (*β* = 0.6*β*_0_) in Fig. 6. Note that, although we chose different introducing date, we run the same transmission period of 500 days. From Fig. 6(a) and 6(c), we can see that the later of the introducing date, the time length that the outbreak takes to peak is shorter. Basically, the reason is that the reproduction number at the initial stage for the introducing time of Jan 1, 2022 is higher than those for the introducing time of Nov 1 and Sep 1, 2021, as shown in Fig. 6(b) and 6(d). On the other hand, we observed an interesting phenomenon that when *κ* = 1, the earlier introducing date caused larger outbreak while when *κ* = 2, the later introducing date caused larger outbreak. Therefore, the peak value of the outbreak is non-monotonous with respect to the introducing time, which is dependent on the ADE effect. Without ADE (i.e. *κ* = 1), a faster growth rate (i.e. the higher effective reproduction number initially) lead to smaller outbreak, as shown in Fig. 6(a) and 6(b). However, when *κ* = 2, a faster infection process means that more population gained the immunity and then lost their immunity, consequently, more susceptible population with ADE effect can be reinfected with a higher peak value (Fig. 6(c) and 6(d)). Therefore, the introducing time can also have a great impact of the transmission dynamics of COVID-19 in the presence of immunity waning.

**Figure 6:**
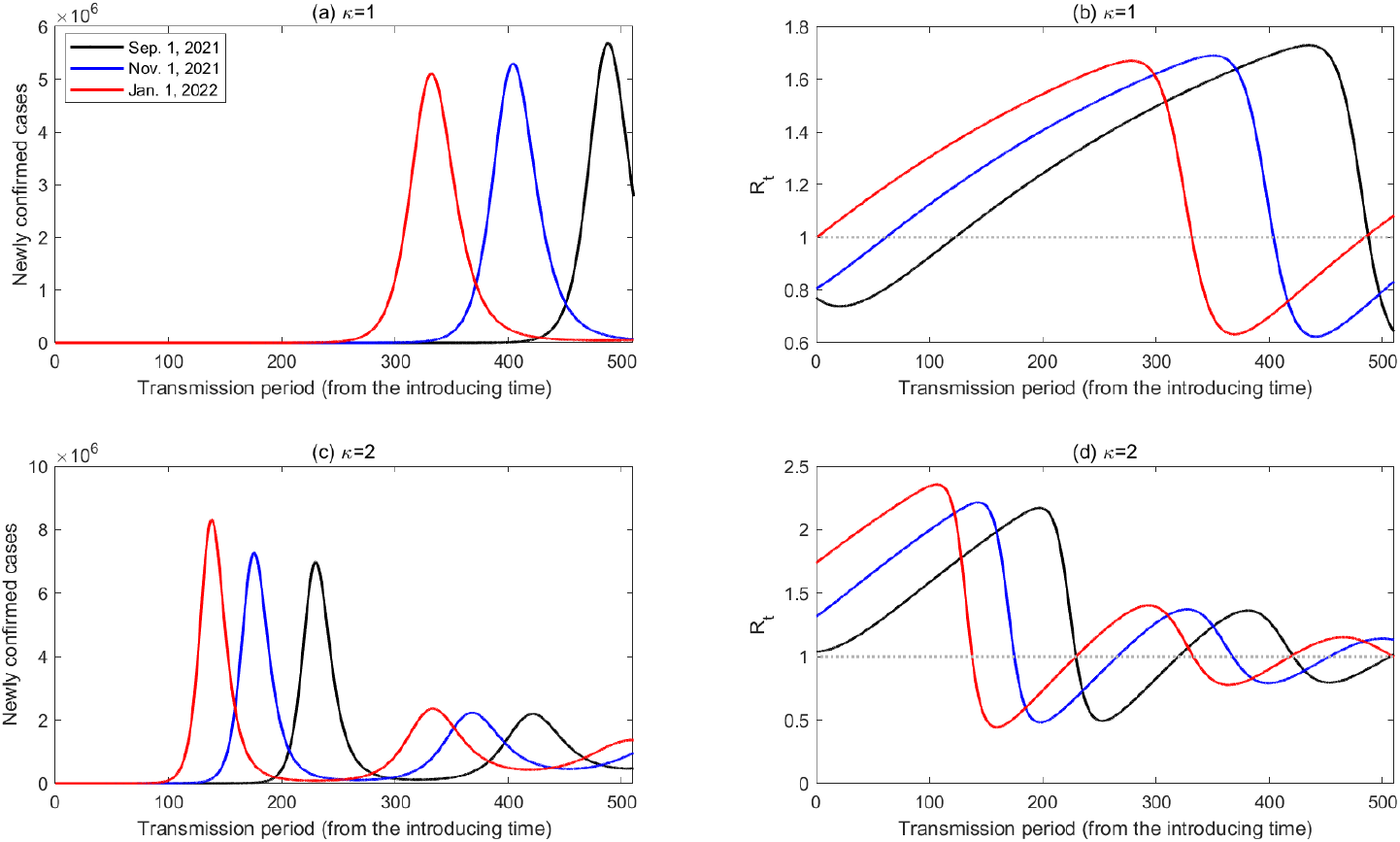
Effect of the ADE and different introducing date on the number of newly confirmed cases and the effective reproduction number during the transmission period.

From the above analysis, we can see that the introducing time can have a great impact on the initial reproduction number because of the immunity waning. Note that, before the introducing date, there is no infected case, hence no transmission in the population. Therefore, the changing of the initial reproduction number totally depends on the immunity waning dynamics without the transmission dynamics. For this reason, we define a new reproduction number as *R*_*s*_ = *R*(*s*), which denotes the reproduction number at the introducing time *s*. In Fig. 7(a) and 7(b), we plotted changing curves of *R*_*s*_ by choosing different transmission rate *β*, ADE factor *κ* and waning rate *ω*. We can easily see from Fig. 7(a) and 7(b) that *R*_*s*_ is increasing over time because of the immunity waning. On the other hand, with a higher transmission ability or ADE degree or immunity waning rate, the reproduction number at the introducing time is always greater, as shown in Fig. 7(a) and (b). By plotting the partial rank correlation coefficients (PRCCs) [38], we also conducted a sensitivity analysis of *R*_*s*_ with resect to the transmission related parameters (*β, ρ, δ*_*I*_, *θ, γ*_*I*_, *γ*_*A*_) and vaccination related parameters 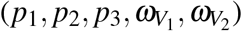 and the ADE factor *κ* over time, as shown in Fig. 7 (c). As a results, we found that the transmission rate always have the most significant effect and is positive related to *R*_*s*_. *κ* does not dominant before June 13, 2021, which means that ADE almost has no effect in the early stage of the vaccination program, mainly due to the high effectiveness of the vaccine and majority of the population had not been vaccinated. However, *κ* positively affects the reproduction number significantly after June 13, 2021, attributing to the fact that people vaccinated may lose the immunity and has the ADE after a period of time. In particular, the PRCCs with respect to the vaccination parameters and the ADE factor (Fig.7)(d) showed that the immunity waning rate 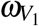 of those received the first dose is also positive related to *R*_*s*_, which is high in the initial stage of the vaccination program. The PRCC of the immunity waning rate 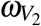 is high in the middle stage of the vaccination program, and the PRCC of the efficacy of two-doses *p*_2_ becomes high in the late stage, illustrating the evolution of the vaccination and immunity waning dynamics.

**Figure 7:**
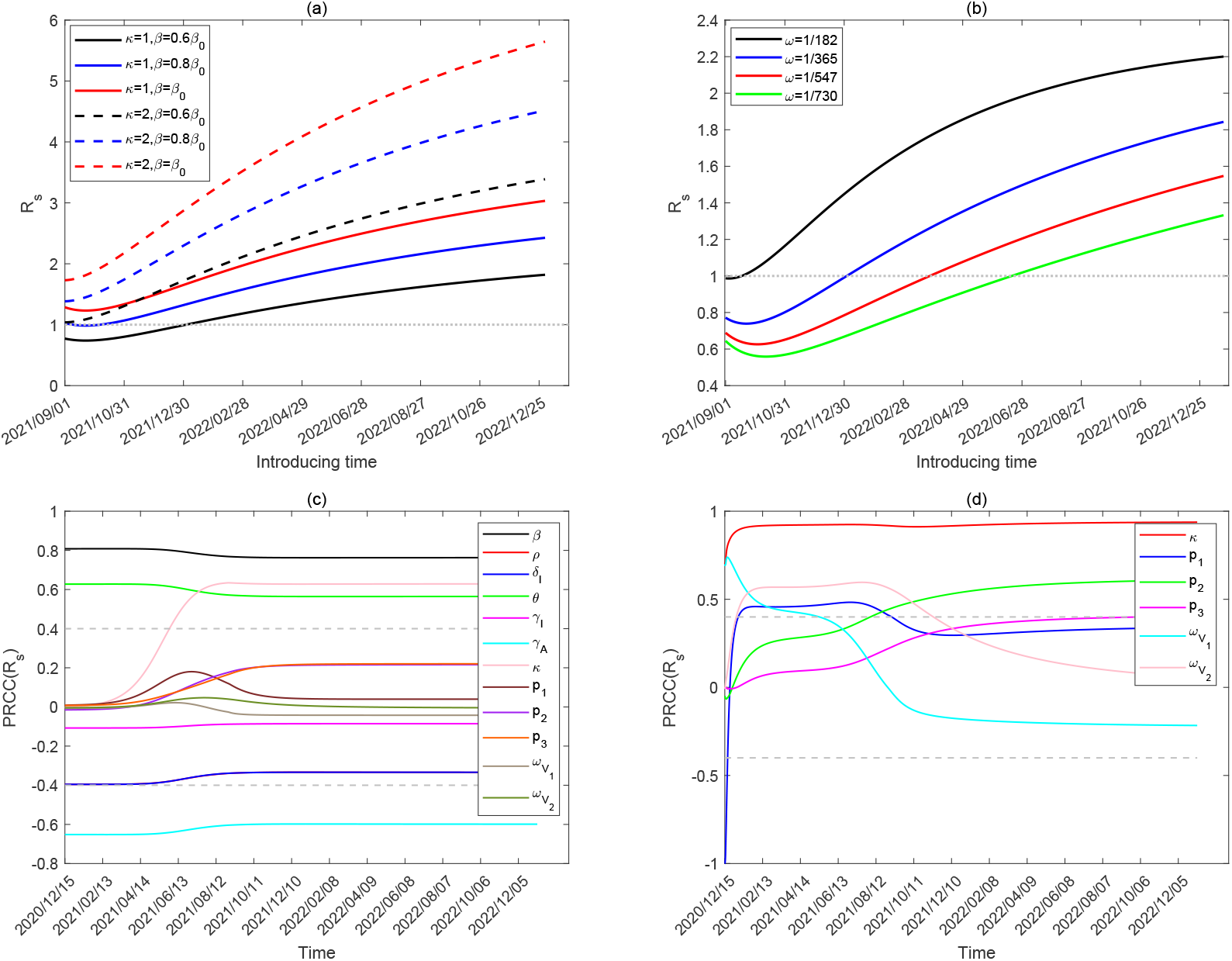
(a) Effect of *β* and *κ* on the initial reproduction number *R*_*s*_. (b) Effect of *ω* on the initial reproduction number *R*_*s*_. (c) Partial rank correlation coefficients (PRCCs) of the initial reproduction number *R*_*s*_ at different introducing time *s* with respect to parameters related to the disease transmission dynamics (*β, ρ, δ*_*I*_, *θ, γ*_*I*_, *γ*_*A*_) and parameters related to the vaccination program 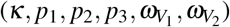. (d) Partial rank correlation coefficients (PRCCs) of the initial reproduction number *R*_*s*_ at different introducing time *s* with respect to the ADE factor (*κ*), vaccination efficacy (*p*_1_, *p*_2_, *p*_3_), and vaccination protective period 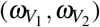. Other parameters are fixed as in Table 1.

In conclusion, we have defined two time-varying reproduction numbers *R*_*t*_ and *R*_*s*_. Here, we tried to combine the two time-varying reproduction number together, and denoted it as *R*(*t, s*), where *s* is the introducing time since the initial time and *t* is the transmission period since the introducing time. Therefore, we have that *R*_*s*_ = *R*(0, *s*) denotes the reproduction number at the introducing time *s* while *R*_*t*_ = *R*(*t*, 0) is the traditional effective reproduction number at time *t*, when the introducing time is taken as the initial time of the epidemic. With this definition, we can easily check the effective reproduction numbers of an epidemic starting at different time. In particular, Fig. 8 show the contour plot of *R*(*t, s*) with respect to different introducing time *s* (taking September 1, 2021 as the initial introducing time) and the transmission period *t* (taking the introducing time as initial time of the transmission process), with the baseline transmission rate *β* = 0.6*β*_0_ and the ADE factor *κ* = 1 and *κ* = 2, respectively. From Fig. 8, we can easily see that *κ* magnify *R*(0, *s*) and makes *R*(*t, s*) tend to be stabilized more quickly. Furthermore, *R*(0, *s*) increases as *s* increases, and *R*(*t, s*) increase first and then fluctuates around the unit with respect to *t* given arbitrary introducing time *s*, which is also observed in Fig. 5 and 6.

**Figure 8:**
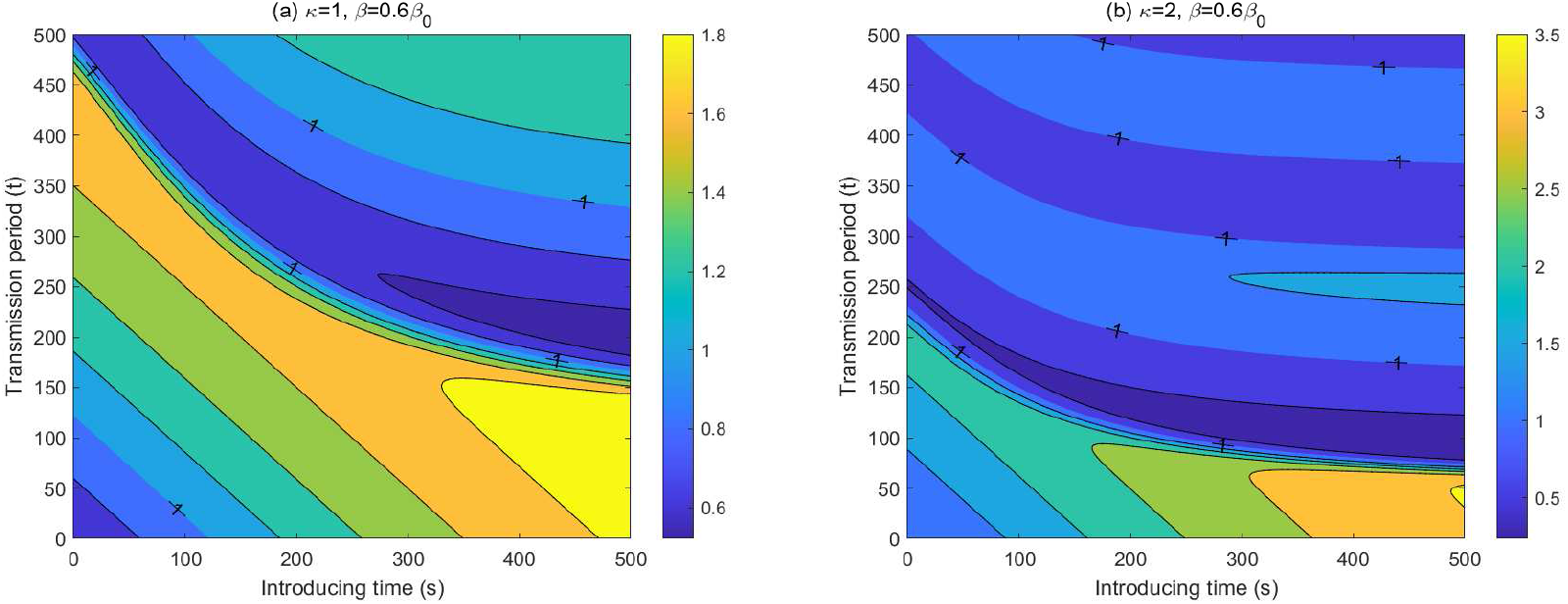
Values of *R*(*t, s*) with different introducing time *s* (taking September 1, 2021 as the initial introducing time) and transmission period *t* (taking the introducing time as initial time of the transmission process).

### 3.3. Protective period evaluation and analysis

Usually, the effective reproduction number is taken as the only risk index to show if the epidemic is under control or not. That is because the new infection will decrease when the effective reproduction number is less than the unit. However, as we illustrated in [39], that the effective reproduction number is less than 1 does not mean that the epidemic is totally under control or we can finally achieve the goal of no newly reported case. It will take a long period to achieve the goal of no newly reported cases. During this period, because of the existence of the source of infection, the COVID-19 epidemic can easily boost once the normalized control interventions are released. Similarly, we cannot say that it will be out of control when the effective reproduction number is greater than the unit when several infected cases are introduced into the community. Therefore, we provide a new definition to represent when an emerging outbreak of COVID-19 can be under control or of a low level of risk.

For any given number of infected cases (*I*_0_) being introduced into the community, it’s reasonable to say that if the newly reported cases is always lower than *I*_0_ (or higher than *I*_0_ but lower than 5*I*_0_) in the next 30 days, it’s of a low (or medium) level of risk in terms of the intraducing time. Otherwise, we say that it’s of a high level of risk. Based on the definition, we can further define a critical time *T*_1_ before which the new epidemic is in a low level of risk if several cases are introduced into the community. That is, *T*_1_ is the first time at which several cases are introduced into the community, and the newly reported cases will reach *I*_0_ at the 30th day. Similarly, we can define the critical time *T*_2_, at which several cases are introduced into the community, and the newly reported cases will reach 5*I*_0_ at the 30th day. Consequently, for the introducing time before *T*_1_ (or between *T*_1_ and *T*_2_, or after *T*_2_), the epidemic is in a low (or medium or high) level of risk. Note that, we observed an interesting phenomenon that for any given number of infected cases *I*_0_ introduced at time *s*, the time length it takes for the number of newly confirmed cases increasing to *kI*_0_ (*k* ≥ 1) is independent of the value of *I*_0_, see a theoretical illustration in SI. This means that the critical time *T*_1_ and *T*_2_ is also independent of the number of cases introduced into the community.

In Fig. 9, we plotted the time length that it takes for the number of newly confirmed cases to reach *I*_0_ or 5*I*_0_ since the introducing time, respectively. Thus, the intersection points of the curves and the horizontal dash line are actually the critical time *T*_1_ or *T*_2_. It follows from Fig. 9(a) and 9(b) that the later introducing of infected cases, the shorter time length it takes for the number of newly confirmed cases increasing to *I*_0_ or 5*I*_0_. Comparing the dash or solid curves with different colors in Fig. 9(a) or 9(b), we find that the increases of the transmission rate and the ADE degree will bring forward the critical time *T*_1_ and *T*_2_, consequently shorten the time period during which the epidemics is of low risk, and bring forward the time after which the epidemic is of high risk by introducing several infected cases. In particular, in the baseline situation, i.e. *κ* = 1, *β* = 0.6*β*_0_, introducing infected cases before the end of the year 2022 would not lead to a large outbreak quickly (in a low level of risk). That is, in this situation, the emerging outbreak by introducing infected cases is always in a low risk till December 31, 2022. When the transmission rate increases to 0.8*β*_0_ or *β*_0_, corresponding to the release of normalized control interventions, the emerging outbreak is in the low level of risk, by the cases introduced before April 28, 2022 or January 22, 2022, respectively.

**Figure 9:**
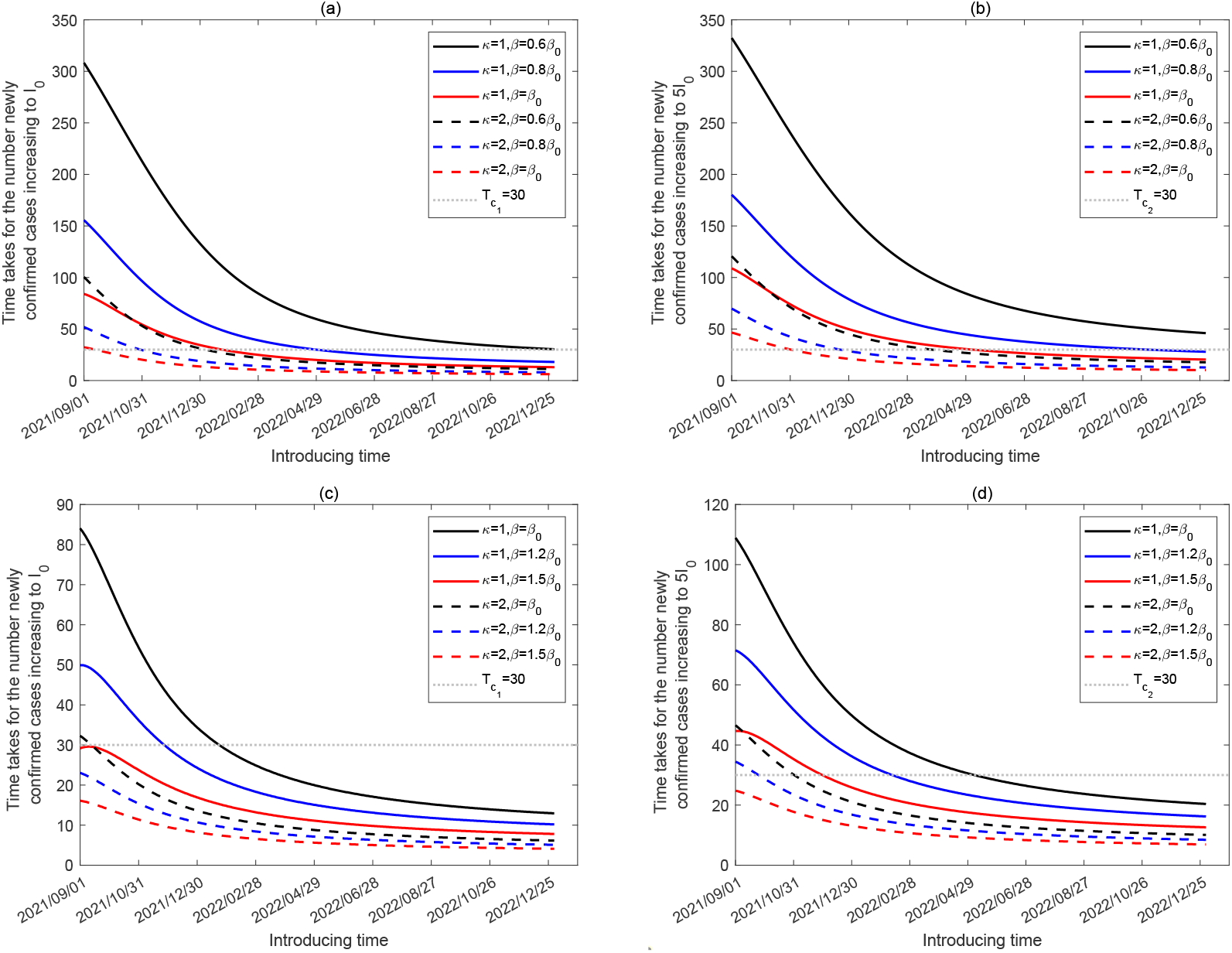
Length of time it takes for the newly confirmed cases increasing to *I*_0_ and 5*I*_0_, respectively, when introducing infected cases at different times for different *β* and *κ*.

In addition, further considering the higher transmission ability of the SARS-CoV-2 variants, we plotted the time length it takes for the confirmed cases to reach *I*_0_ or 5*I*_0_ since the introducing time in Fig. 9(c) and (d), by choosing the transmission rate as 1.2*β*_0_ and 1.5*β*_0_. It follows from Fig. 9(c) that when *κ* = 2, *β* = 1.2*β*_0_ or *κ* = 2, *β* = 1.5*β*_0_, the curves are always below the horizontal dash line since September 1, 2021. This means that if the ADE is possible, the emerging outbreak of SARS-CoV-2 variants can not be in a low level of risk by introducing new infected cases since September 1, 2021, and it’s in a high level of risk since the initial time we considered by introducing infected cases when *κ* = 2, *β* = 1.5*β*_0_ (Fig. 9(d)). On the other hand, if we fix *β* = 0.6*β*_0_ but assume that that the ADE is possible by letting *κ* = 2, the critical time *T*_1_ and *T*_2_ becomes January 4, 2022, and March 25, 2022, respectively, as shown in Fig. 9(a),(b) and Table 2. To summary, Table 2 lists the critical time *T*_1_ and *T*_2_ under the different situations with different transmission rate and ADE degree.

**Table 2:** The impact of the transmission rate and ADE factor on the time period of the low, medium and high risk levels by introducing infected cases.

| Parameters | Low risk period | Medium risk period | High risk period |
| --- | --- | --- | --- |
| $\kappa = 1, \beta = 0.6\beta_0$ | before 2023/01/18 | - | - |
| $\kappa = 1, \beta = 0.8\beta_0$ | before 2022/04/28 | 2022/04/28 - 2022/11/01 | after 2022/11/01 |
| $\kappa = 1, \beta = \beta_0$ | before 2022/01/22 | 2022/01/22 - 2022/05/05 | after 2022/05/05 |
| $\kappa = 1, \beta = 1.2\beta_0$ | before 2021/11/26 | 2021/11/26-2022/02/10 | after 2022/02/10 |
| $\kappa = 1, \beta = 1.5\beta_0$ | - | before 2021/11/29 | after 2021/11/29 |
| $\kappa = 2, \beta = 0.6\beta_0$ | before 2022/01/04 | 2022/01/04-2022/03/25 | after 2022/03/25 |
| $\kappa = 2, \beta = 0.8\beta_0$ | before 2021/10/30 | 2021/10/31-2021/12/21 | after 2021/12/21 |
| $\kappa = 2, \beta = \beta_0$ | before 2021/09/13 | 2021/09/13-2021/11/01 | after 2021/11/01 |
| $\kappa = 2, \beta = 1.2\beta_0$ | - | before 2021/09/24 | after 2021/09/24 |
| $\kappa = 2, \beta = 1.5\beta_0$ | - | - | after 2021/09/01 |

In addition, we showed the contour plots of *T*_1_ and *T*_2_, respectively, by regarding September 1, 2021 as the initial time 0 in Fig. 10, with respect to the transmission rate *β* and the ADE factor *κ* (Fig. 10(a) and (b)), and with respect to the transmission rate *β* and the waning rate *ω* (Fig. 10(c) and (d)). From Fig. 10 (a) and (b) we can see that *T*_1_ and *T*_2_ decreases with the increase of *β* and *κ*, meaning that the low risk period is shorten and the high risk time is brought forward. *T*_2_ is certainly greater than *T*_1_ while for variants with high transmission ability and strong ADE degree, it’s hard to have a period during which introducing infected cases is of low risk. However, with strict normalized control interventions (small transmission rate, for example *β* = 0.4*β*_0_), even if ADE is slightly feasible (*κ* varies from 1 to 1.5), the emerging outbreak of introducing infected cases would maintain in a low risk till December 31, 2022. From Fig. 10 (c) and (d), we can see that the increasing of the waning rate *ω* also leads to the decrease of *T*_1_ and *T*_2_. When the transmission rate increases to 0.8*β*_0_, a reduced immunity waning rate *ω* = 1*/*180, can also ensure the emerging outbreak in a low risk by introducing infected cases by the end of the year 2022. However, when the transmission rate is small enough, corresponding to the strict normalized control strategies, the emerging outbreak of introducing infected cases is always maintained in a low risk till December 31, 2022, regardless of the waning rate. Both contour plots illustrated that strengthening normalized control interventions is quite efficient in protecting the community from the rapidly outbreak induced by the imported infected cases.

**Figure 10:**
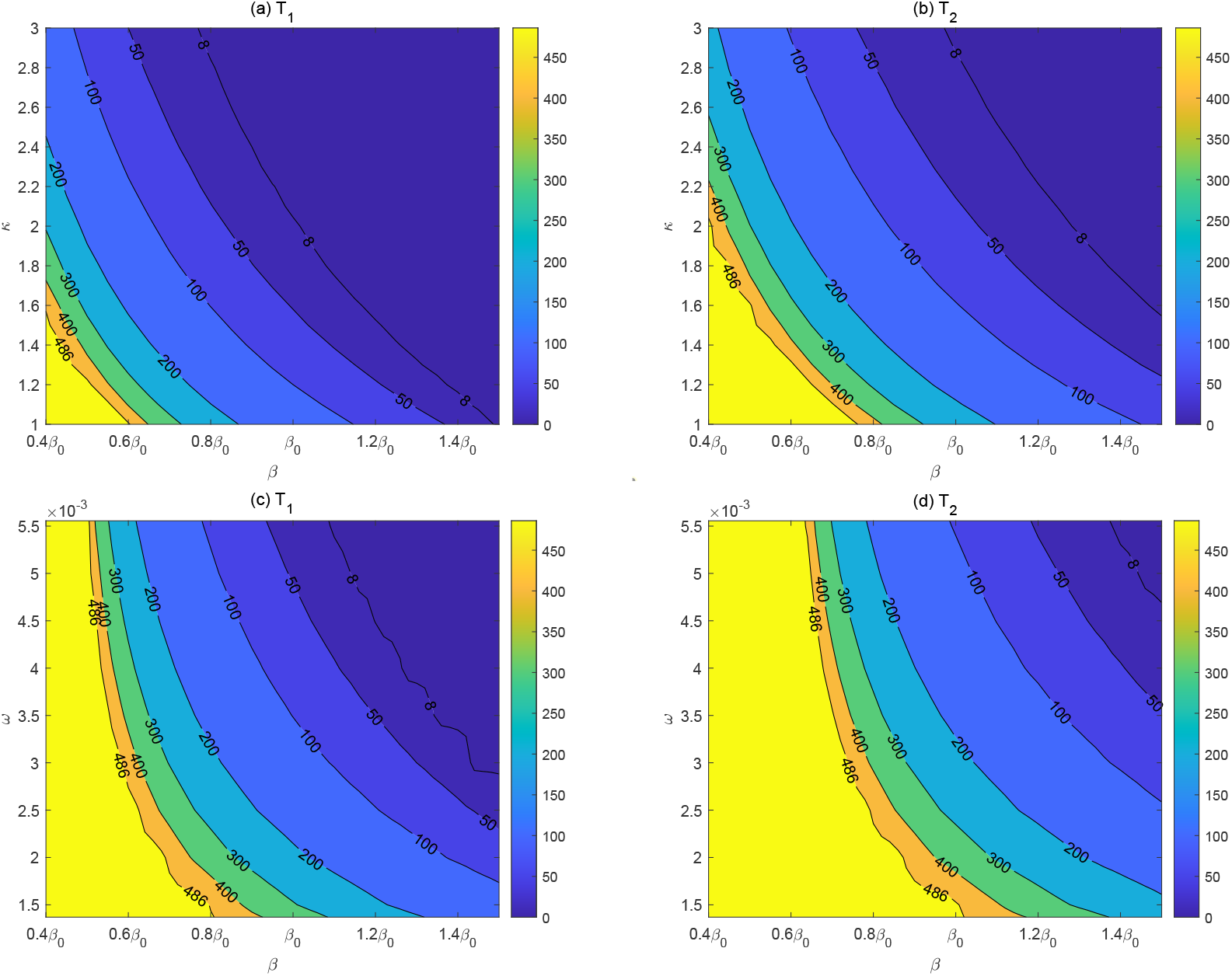
Contour plots of *T*_1_ and *T*_2_ with respect to *β* and *κ, β* and *ω*, by taking September 1, 2021 as the initial time.

## 4. Discussion

Creating the herd immunity by using COVID-19 vaccines in the population has been taken as the only solution to stop the COVID-19 pandemic for a long time. Especially for the counties or territories experienced multiple epidemic waves even with various non-pharmaceutic interventions (NPIs). We should be sure that vaccines do work to slow down the epidemics. However, immunity waning and ADE may destroy the hope of building the herd immunity to finally eradiate SARS-CoV-2. Therefore, it’s urgent and critical to evaluate how long the current vaccination program can protect the country in a low level of risk in the presence of immunity waning, and ADE. This can provide important decision making-basis for the decision makers to determine when the catch-up vaccination program should be launched.

In the current study, we take mainland China as the example to illustrate the possible transmission dynamics of COVID-19 with the current vaccination program. Incorporating the immunity waning dynamics and ADE effect, we developed a new mathematical model. The proposed model was calibrated by using the data of the COVID-19 epidemic in 2020 in mainland China, and the vaccination data from Dec. 15, 2020 to Jun 29, 2021. The estimation showed that the cumulative population with at least one dose reached 56.4% while the population with two doses reached 32.02% on June 29, 2021 (the last data point). A prediction indicated that the vaccination coverage with at least one dose would reach 95.87% and the proportion with two-doses would reach 77.92% on August 31, 2021. Therefore, the vaccination coverage has already reached a very high level in China, reflecting a spectrum of immunity.

We initially tested if we can return to the pre-COVID-19 pandemic era or not by the mass vaccination program only. That is, we showed if the emerging epidemics, by introducing several new cases into communities, can be under control or not without any other NPIs (i.e. *β* is set to be *β*_0_). The answer is definitely no in the presence of immunity waning. We found that the daily reported cases can grow exponentially in a short period since the introducing time, and peak at a huge number. This is directly due to the immunity waning, and a large proportion of vaccinated individuals become susceptible again. We can intuitively see the reason from the newly defined reproduction number *R*_*s*_, i.e. the reproduction number at the introducing time. *R*_*s*_ increases and exceeds the threshold of unit as the introducing time postpones, attributing to the dynamic of immunity waning in the population. That’s also the reason why the introducing time of infected cases has a great influence on the transmission dynamics of COVID-19 epidemics (6). Generally, the later of the introducing time, the shorter period that the daily reported cases take to peak. This indicates that the faster implication of interventions should be considered when the introducing time is long from the stopping time of the vaccination program. We observed an interesting phenomenon that the peak value of the outbreak is non-monotonous with respect to the introducing time.

We also observed from Fig. 5 and 6 that intermittent outbreaks of COVID-19 can occur. Initially, the immunity waning leads to the breakthrough of the herd immunity, consequently, introducing new infected cases can result in a large outbreak. In return, the repeated outbreaks can further boost the herd immunity in the population level and drive the decline of the effective reproduction number, subsequently drive the decline of the epidemics. In conclusion, a loop of immunity waning and immunity boosting in the population induced the intermittent epidemic. Furthermore, it should be mentioned that the amplitudes of the subsequential outbreaks are decreasing over time. This means that a large proportion of the population will be protected sufficiently after several outbreaks. This result implies that boosting the immunity by a booster injection of vaccine in the population may be helpful in mitigating the possible outbreaks. The optimized boosting program should be further studied.

Although the reproduction number can be an important index to represent if the number of newly infected cases is increasing or not, it doesn’t mean that the epidemic will be out of control when the effective reproductions number is greater than the unit, as it can be just slightly greater than the unit in a relatively long period. Therefore, from another point of view, we tried to find a new index to represent whether the emerging epidemic is under control. That is, for any given number of cases (*I*_0_) introduced into the community, we say if the newly reported cases is always lower than *I*_0_ in the next 30 days, it’s of a low level of risk in terms of the introducing time. With our definition, we found that any emerging epidemic of COVID-19 by introducing infected cases before Januray 22, 2022 can be in a low level of risk. As we can see, it’s very short for the protective period of the vaccination program. For this reason, we assumed 50% populations to maintain a normalized control interventions by wearing masks, correspondingly, the transmission rate is set to be 0.6*β*_0_. The results showed that the vaccination program incorporating a normalized interventions can prolong the protective period till January 18, 2023.

ADE is thought to be a big trouble in the development and use of COVID-19 vaccines. We also quantitatively evaluated the impact of ADE on the transmission dynamic of COVID-19 with the implementation of the mass vaccination program. The intuitive results are that ADE can bring forward the peak time of an outbreak and greatly increase the peak value of the daily reported cases. Furthermore, ADE can also increase the frequency of the intermittent outbreaks. In our definition, with a normalized control intervention, ADE can bring forward the critical time of escaping the low risk almost one year (i.e. from January 18, 2023 to January 4, 2022), as listed in Table 2. Considering the higher transmission ability of the SARS-CoV-2 variants, we obtained the similar results as those of ADE. Apparently, ADE and the emerging of new variants have made the control of COVID-19 epidemics even more difficult.

## 5. Conclusion

Utilizing mathematical model, this study focused on investigating the resurgence risk of COVID-19 after the mass vaccination program in China in the presence of immunity waning and ADE. The vaccination coverage is projected to be very high till Aug. 31, 2021, which would almost reach the requested critical level of herd immunity. However, the herd immunity can be easily breakthrough by immunity waning. Therefore, we suggest to maintain a normalized control interventions of wearing masks in the long future even with the mass vaccination program. By defining the risk level of an emerging outbreak, our results showed that the current vaccination program incorporating the normalized interventions can protect China in a low level of resurgence risk till 2023/01/18. However, the emerging evidence of the existence of ADE and the SARS-CoV-2 variants with higher transmission ability have made the situation worse. Therefore, we should get ready for a long struggle with COVID-19, and should not totally rely on COVID-19 vaccines.

It’s worth mentioning that boosting the immunity in the population might be able to mitigate the emerging outbreaks. Maintaining the normalized NPIs and booster injection of vaccination periodically could be a good choice in combating with COVID-19 in the long future. How to optimize the periodic vaccination program incorporating the implementation of NPIs is of great significance, which falls within the scope of our further work.

## Data Availability

The epidemic data is public available.

## Acknowledgements

The authors are supported by the National Natural Science Foundation of China (grant numbers: 12031010(ST),11631012(YX),12001349(WZ)), the Postdoctoral Research Foundation of China (grant number: 2019M663611 (WZ)), and the Young Talent Support Plan of Xian Jiaotong University (BT).

## Supplementary Information

### 1 The model

#### Full structure model

The full modelling framework includes the infection and transmission dynamic of COVID-19 and the two-doses vaccination program and immunity waning dynamic.

##### Transmission progression

We used an *SEIARS* model structure to describe the transmission dynamics of COVID-19. That is, according to the epidemic status of COVID-19 infections, the total population is divided into susceptible (*S*), exposed (*E*), symptomatic infected (*I*), asymptomatic infected (*A*), recovered (*R*) classes [2, 3, 4]. The susceptible population will enter into the exposed class (*E*) once they are infected by symptomatic or asymptomatic infected population. The exposed individuals move to *I* or *A* at a rate of *σ*. And we assume that the probability of showing symptoms is *ρ*. The recovery rates of *I* and *A* are set to be *γ*_*I*_ and *γ*_*A*_, respectively. Given the immunity waning, the recovered population can become susceptible again. The transmission diagram with the mass vaccination program is shown in Fig. 1.

##### Vaccination program for susceptible population

Susceptible population (*S*), will be firstly vaccinated by one-dose at a rate *v*_1_. The effective protection rate by one-dose is *p*_1_, hence, the part *p*_1_*v*_1_ can be effectively protected and temporarily immune to COVID-19, and the class is denoted by *V*_1_. The rest part ((1 − *p*_1_)*v*_1_) remains in the susceptible class, which is denoted by 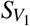. The population vaccinated by one-dose will be further vaccinated by a second dose after a pre-set period since the first dose. Similarly, we set a protection rate of the second dose as *p*_2_. Once vaccinated by the second dose, individuals will either move to the class effectively protected (denoted by *V*_2_), or to the class susceptible to COVID-19 (denoted by 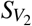). We assume that the population effectively protected by one-dose (the persons in *V*_1_), will all move to *V*_2_ when they receive the second dose. It should be mentioned that the population vaccinated but not immune to COVID-19 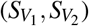 can be infected. Then according to the epidemic status, we further have the classes of *E*_*j*_, *I*_*j*_, *A*_*j*_, *R*_*j*_, *j* ∈ {*V*_1_,*V*_2_} as shown in Fig. 1.

##### Immunity waning and ADE effects

As we mentioned in the introduction, lots of evidence suggest that the neutralizing antibodies decay significantly since the onset of symptoms of COVID-19 patients, indicating the existence of immunity waning. Given the immunity waning, we assume that both the recovered and effectively vaccinated population can temporarily immune to COVID-19, and will return back to susceptible classes. In particularly, we assume that the individuals in *V*_1_ will move to 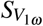 due to the waning of immunity, and the population in 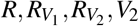 will move to 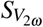. Correspondingly, the rate of immunity waning are denoted by 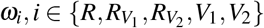. On the other hand, as the decay of the neutralizing antibodies, the binding antibodies can dominant the immune response, which will enhance the infectivity of the virus (i.e. ADE effects). That is, compared with other susceptible population, the susceptibility of 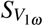 and 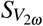 is much higher as the pre-existing of immunity. Here, we denote *κ* as the modification factor of the susceptibility of the individuals who have lost their immunity.

Based on the above assumptions, the model equations can be written as:

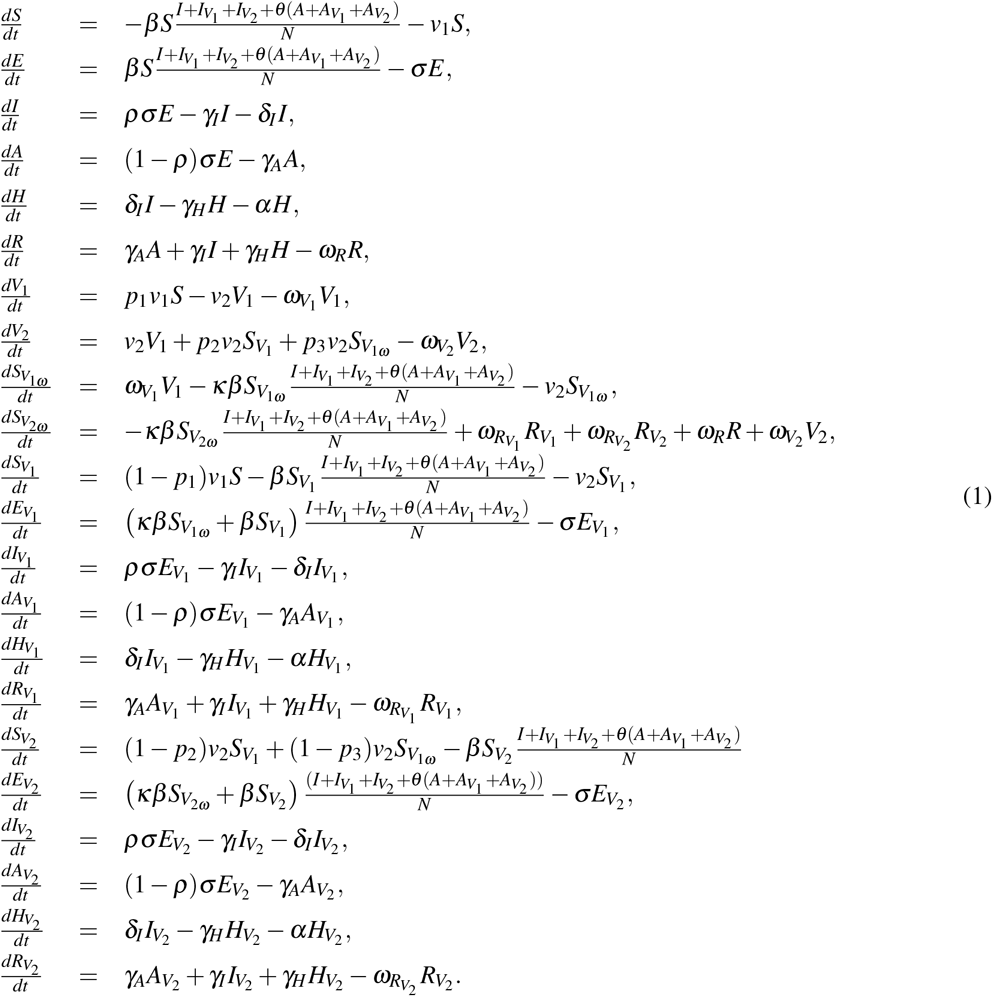

Here, *N* is the whole population in the considered region. The equations for *S, E, I, A, H, R* represent the infection and transmission dynamic in the non-vaccinated population, the equations for 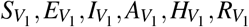represent the infection and transmission dynamic in the population vaccinated with the first dose vaccine, the equations for 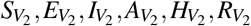 represent the infection and transmission dynamic in the population vaccinated with two doses vaccine. *V*_1_ and *V*_2_ are the population effectively protected by the first dose and two doses vaccine, respectively, who would not evolve in the transmission dynamic but will lose the immunity with rate 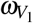 and 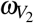, respectively. 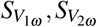 are the population lost the immunity and are more susceptible to the infection due to ADE effect. The detailed definitions of all the parameters are listed in Table 1.

Using the next generation matrix method, we can firstly calculate the basic reproduction number [1]. Then subsisting the time-varying parameters and variables into the basic reproduction number, we can obtain the effective reproduction number of model (1), which is given by

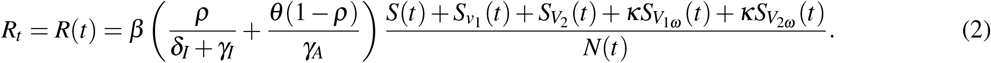

#### Transmission dynamic model without vaccination dynamic

Note that, during the outbreak of COVID-19 in mainland China in 2020, there is no vaccination, consequently, model (1) can be reduced to

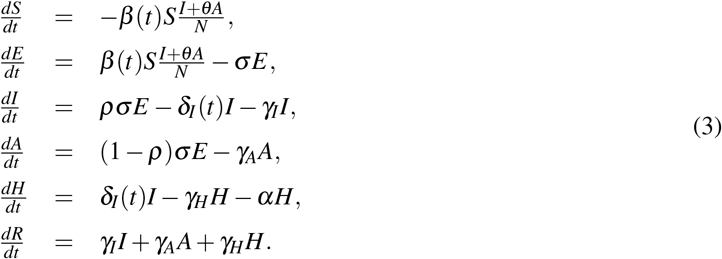

Here, considering the continuously enhanced control interventions by the government, we introduced two time-dependent parameters into the system. In detail, the transmission rate *β* is set to be a decreasing function of time *t* with the following form

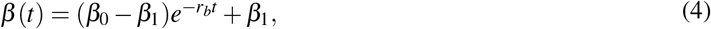

where *β*_0_ is the initial transmission rate, *β*_1_ is the minimum transmission rate, and *r*_*b*_ is the exponential decreasing rate of the transmission rate. Similarly, we set the diagnose rate is a increasing function of time *t* with the following form

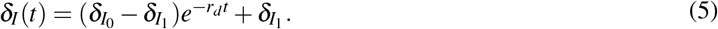

where 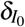 is the initial diagnose rate, 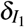 is the maximum diagnose rate, and *r*_*d*_ is the exponential increasing rate of the diagnose rate. *t*_0_ corresponds to January 23, 2020.

Model (3) with (4) and (5) can be used to describe the transmission dynamic of the COVID-19 epidemics in mainland China in 2020, with the gradually improved and enhanced non-pharmaceutic interventions (NPIs), and without vaccination. Therefore, model (3) can be used to fit the epidemic data of COVID-19 epidemic in mainland China in 2020, to obtain the values of parameters related to the disease transmission.

#### Vaccination dynamic model without transmission dynamic

Since the initiation of the vaccination program, it has already taken more than 9 months to vaccinate against COVID-19 in mainland China. Therefore, the vaccination program is also a dynamic process. To model the vaccination dynamic without the transmission dynamic, a three-compartment model reduced from model (1) was derived. The three compartments are the population without vaccination (*S*), the population got the first vaccine dose (*S*_1_) and the population who have received the second vaccine dose (*S*_2_), respectively. Then the model is given by:

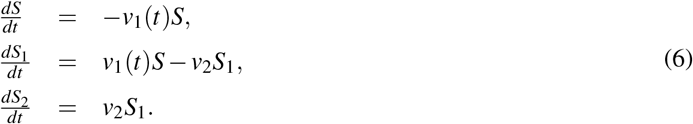

where *v*_1_(*t*) is the time-dependent vaccination rate, as it should be small initially since the availability of the number of vaccine doses was limited at the beginning, and then exponentially increased as the production of COVID-19 vaccines was accelerated, and finally it could plateau to a constant level depending on the daily vaccination capacity. Thus, we assume *v*_1_(*t*) as a logistic increasing function of time *t* with the following form:

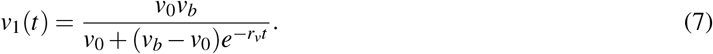

with *t* corresponds to the time when the initial time is assumed to be December 15, 2020. As a result, model (6) can be used to fit the vaccination data in mainland China, to obtain the values of parameters related to the vaccination program.

#### Immunity waning dynamic model without transmission dynamic

Note that immunity waning makes the people who have gained the immunity after vaccination become susceptible again, thus based on model (1) and incorporating the natural immunity waning into model (6), we can obtain the following system:

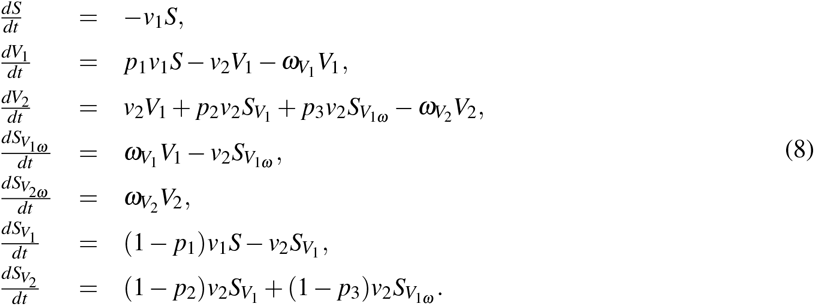

Then, model (8) combined the vaccination dynamic and the nature immunity wanning dynamic, but without the transmission dynamic of COVID-19. That is, model (8) can be used to simulate the vaccination dynamics and the immunity waning dynamic during the pre-epidemic period.

### 2 Theoretical illustration of *T*_1_ and *T*_2_ remain constant with different *I*_0_

We will briefly illustrate that the time length it takes for the number of newly confirmed cases reaching the level of initial introducing infected cases is independent of the initial value of the introducing infected cases, by using a simple *SEIAHR* model in the following.

Assume that several infected cases (*I*_0_) are introduced at time *t*_0_ in a fully susceptible population, then the system

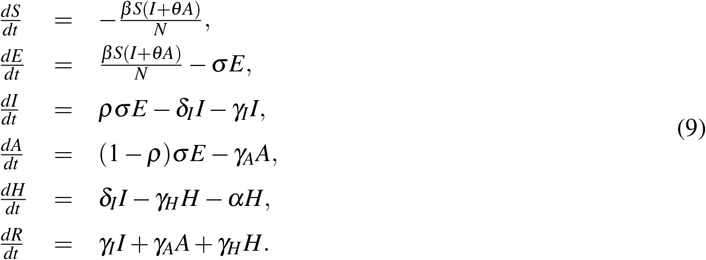

has the initial condition (*S*(*t*_0_), *E*(*t*_0_), *I*(*t*_0_), *A*(*t*_0_), *H*(*t*_0_), *R*(*t*_0_)) = (*S*_0_, 0, *I*_0_, 0, 0, 0) at the initial time *t*_0_, and *I*_0_ ≪ *N*_0_. Thus it is reasonable to assume 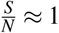 in the initial stage of the disease transmission, then by omitting the equation of *S* and *R*, the model can be reduced to

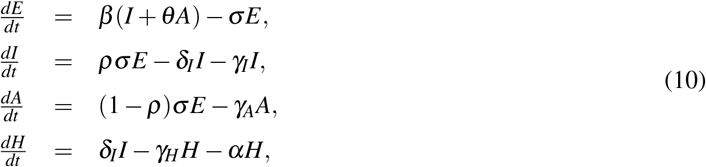

with initial condition (*E*(*t*_0_), *I*(*t*_0_), *A*(*t*_0_), *H*(*t*_0_)) = (0, *I*_0_, 0, 0). Denote 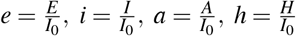, system (10) is equivalent to the following system

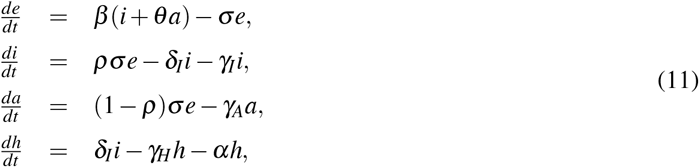

with initial value (0, 1, 0, 0). Thus the solution of system (11) is independent of the value of *I*_0_ and is unique. Thus *t*_1_ is constant regardless the value of *I*_0_, where *i*(*t*_1_) = 1*/δ*, namely *δ I*(*t*_1_) = *I*_0_. Thus illustrating that the time it takes for the newly confirmed cases *δI* increases to the initial value *I*_0_ is independent of *I*_0_.

## Notes

### Competing Interest Statement

The authors have declared no competing interest.

### Funding Statement

The study is supported by the National Natural Science
Foundation of China (grant numbers: 12031010(ST),11631012(YX),12001349(WZ)), the Postdoctoral Research Foundation of China (grant number: 2019M663611 (WZ)), and the Young Talent Support Plan of Xi'an Jiaotong University (BT).

### Author Declarations

The data were released and analysed anonymously.

